# Association between combined sewer overflow events and gastrointestinal illness in Massachusetts municipalities with and without river-sourced drinking water, 2014-2019

**DOI:** 10.1101/2023.10.25.23297573

**Authors:** Beth M. Haley, Yuantong Sun, Jyotsna S. Jagai, Jessica H. Leibler, Robinson Fulweiler, Jacqueline Ashmore, Gregory A. Wellenius, Wendy Heiger-Bernays

## Abstract

**Background:** Combined sewer overflow (CSO) events release untreated wastewater into surface waterbodies during heavy precipitation or snowmelt. Combined sewer systems serve approximately 40 million people in the United States, primarily in urban and suburban municipalities in the Midwest and Northeast. Predicted increases in heavy precipitation events driven by climate change underscore the importance of quantifying potential health risks associated with CSO events.

**Objectives:** The aims of this study were: to 1) estimate the association between CSO events (2014-2019) and emergency department (ED) visits for acute gastrointestinal illness (AGI) among Massachusetts municipalities that border a CSO-impacted river, and 2) determine if associations differ by municipal drinking water source.

**Methods:** A case time series design was used to estimate the association between daily cumulative upstream CSO discharge in the prior four days and ED visits for AGI, adjusting for temporal trends, temperature, and precipitation. Associations between CSO events and AGI were also compared by municipal drinking water source (CSO-impacted river vs. other sources).

**Results:** Extreme upstream CSO discharge events (>95th percentile by cumulative volume) were associated with a cumulative risk ratio (CRR) of AGI of 1.22 (95% CI: 1.05, 1.42) over the next four days for all municipalities, and the association was robust after adjusting for precipitation (1.17 [0.98, 1.39]). In municipalities with CSO-impacted drinking water sources, the adjusted association was somewhat less pronounced following 95th percentile CSO events (1.05 [0.82, 1.33]). The adjusted CRR of AGI was 1.62 in all municipalities following 99th percentile CSO events (95% CI: 1.04, 2.51) and not meaningfully different across strata defined by drinking water source.

**Discussion:** In municipalities bordering a CSO-impacted river in Massachusetts, extreme CSO events are associated with higher risk of AGI within four days. The largest CSO events are associated with increased risk of AGI regardless of drinking water source.

## Introduction

In more than 700 municipalities in the United States, combined sewer systems (CSS) collect residential and non-residential sewage, industrial waste, and stormwater that flow together to a wastewater treatment facility. CSS discharge untreated combined wastewater into rivers and lakes when the volume of water in the common collection pipe exceeds treatment capacity, potentially posing risks to the health of communities and ecosystems downstream of discharge points. Discharge events—called combined sewer overflow (CSO) events—are driven by heavy precipitation and/or runoff from snowmelt.^1,2^ Observed and predicted increases in the intensity and frequency of precipitation events due to climate change in the Northeast and Midwest where most CSS are located underscore the importance of understanding and addressing the health risks associated with CSO events.^3–5^

CSO events introduce viral, bacterial, and protozoan pathogens to surface waterbodies, as well as chemical and physical contaminants.^1,2,6–14^ Compared to dry weather conditions, CSO events can lead to one to two orders of magnitude increases in the concentration of waterborne pathogens and fecal indicator organisms in receiving waterbodies.^2,6,15^ Exposure to sewage-associated pathogens can lead to gastrointestinal, respiratory, and soft tissue infections among recreational and drinking water users.^16–21^ The association between fecal contamination of surface waters and gastrointestinal illness is well-documented in settings with modern wastewater infrastructure,^22–25^ as is the relationship between precipitation, which is the primary driver of CSO events, and gastrointestinal illness.^26–28^ However, relatively little research has been conducted to determine the direct association between CSO events and health outcomes.

The few studies that have investigated the relationship between CSO events and health outcomes suggest a link between discharge events and acute gastrointestinal illness (AGI).

Studies of recreational use of CSO-impacted waterbodies suggest that the risk of AGI among swimmers can increase up to five times after a CSO event compared to when no CSO event occurred,^29^ and that the likelihood that recreators will develop AGI can vary widely by activity and level of water contact.^9^ Increases in pediatric cases of AGI following large volume partially treated sewage discharge events in a drinking water source^30^ and increased risk of AGI following extreme precipitation events in a region with a CSO-impaired drinking water source^31^ suggest that an underexplored pathway of exposure may be contaminated drinking water. Importantly, neither of these studies used CSO event data in their analyses, but instead used partially treated sewage discharge event or precipitation data to characterize exposure, respectively.^30,31^ Research has demonstrated elevated rates of AGI following large volume CSO events among residents of Atlanta^32^ and increased odds of AGI among children living near a CSO outfall after any CSO event.^33^ In all studies, the association between discharge events and reported AGI has a lag of two to eight days, which could be due to pathogen travel time in the waterbody, timing of recreational activity, drinking water treatment and distribution, pathogen infection and incubation, and/or decision to seek care.^30–33^.

There remain key gaps in our understanding of the association between CSO events and health, including the robustness of the association, relevant exposure pathways, and the role of precipitation events. Accordingly, we sought to assess the association between CSO events and emergency department (ED) visits for AGI in the Massachusetts municipalities that border the Merrimack River, a drinking water and recreational resource for surrounding communities. We evaluate whether the association between CSO events and AGI differs by municipal drinking water source and after adjusting for precipitation events. We hypothesize that 1) upstream CSO events increase risk of AGI in downstream municipalities after adjusting for precipitation; and 2) the risk of AGI in municipalities that exclusively source their drinking water from the Merrimack River is more strongly associated with upstream CSO discharge than in municipalities with other drinking water sources. To our knowledge, this is the first study that directly investigates the role of drinking water source in the association between CSO event discharge volume and AGI.

## Methods

### Study Site

The Merrimack River flows 115 miles through New Hampshire (NH) and northern Massachusetts (MA) before flowing into the Atlantic Ocean.^34^ At 5,010 square miles, the Merrimack watershed is the fourth largest in New England and home to over 2.6 million people.^34^ Over 500,000 people rely on the Merrimack as a drinking water source, and it is a regional recreational resource for many more.^35^

The main stem of the Merrimack River flows through five of the six CSS municipalities in the watershed (Figure 1). The sixth CSS community in the watershed is located 51 miles upstream from the confluence with the main stem of the Merrimack River along a tributary and not included in this study due to its distance from the main stem of the river. Four municipalities in Massachusetts exclusively source their drinking water from the river, while three intermittently depend on the Merrimack River as a drinking water source. All 17 Massachusetts municipalities that border the Merrimack River are included in this study.

**Figure 1:**
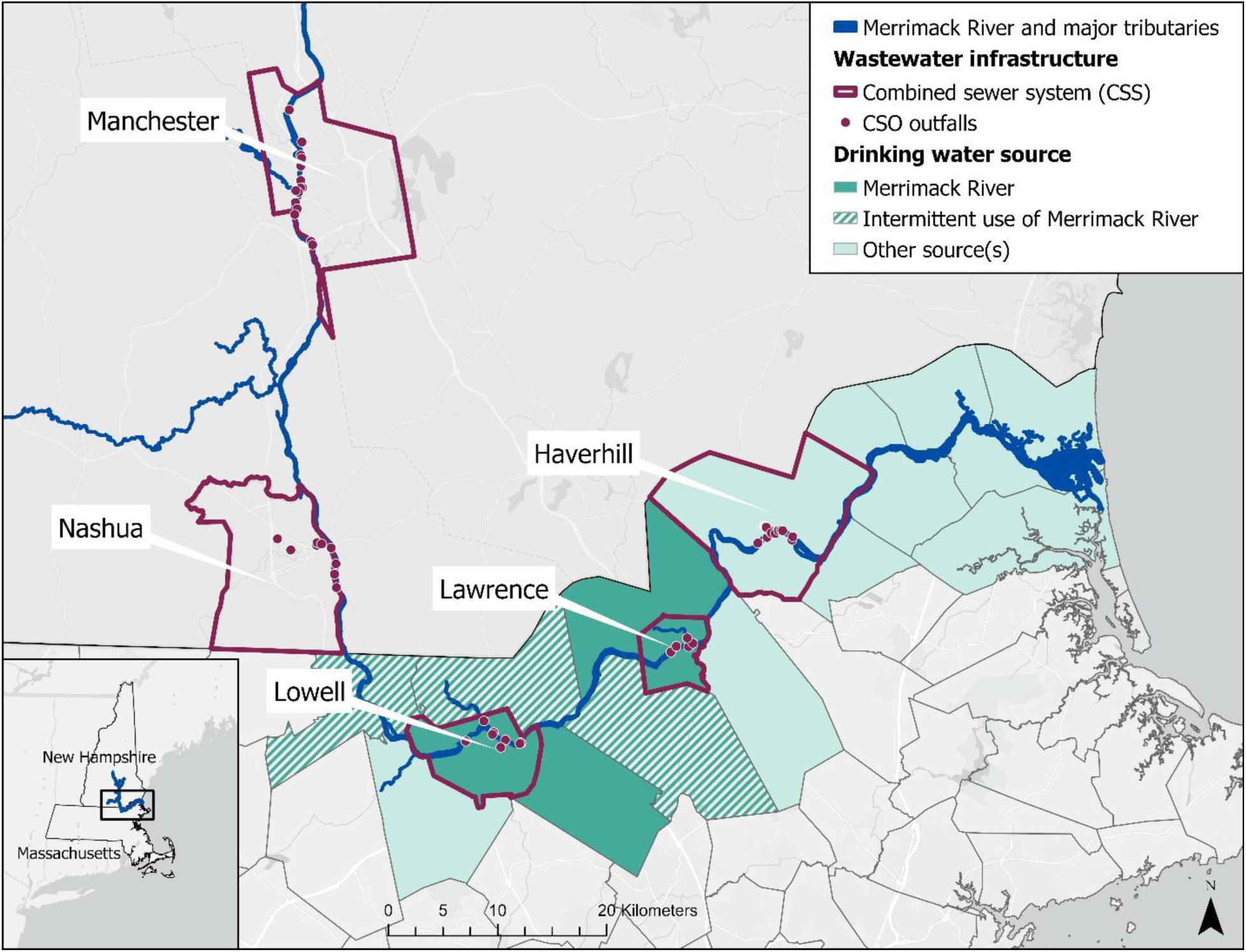
Map of the study area in the lower reaches of the Merrimack River.

### Exposure Data and Classification

US Environmental Protection Agency (EPA) Region 1 and the City of Manchester, NH provided daily CSO event data for the study period. Locations of CSO outfalls are documented and publicly available via an online dashboard.^35^ CSO event data were aggregated by municipality and date, yielding a time series of total daily CSO discharge volume per CSS municipality. Cumulative daily upstream CSO discharge volume was calculated for each study municipality based on which CSS municipalities are located upstream. Figure S1 shows the 17 study municipalities and their spatial relationships to CSS.

Total daily precipitation and mean daily temperature were estimated using PRISM (Parameter-elevation Relationships on Independent Slopes Model) Climate Data.^36^ The centroid for each municipality in the study was used to identify the corresponding 4 km^2^ PRISM grid cell and generate a daily time series of total precipitation and average temperature for each municipality. Cross-correlations were performed to determine how closely and at what point in the time series precipitation and CSO discharge are most correlated.

Study days were categorized in one of the following ways: 1) extreme CSO/precipitation events (≥ 95^th^ percentile events on the basis of cumulative daily upstream CSO discharge or total daily precipitation, respectively), 2) all other CSO/precipitation events (<95^th^ percentile events), or 3) no CSO/precipitation events. Days with no CSO/precipitation events were used as the referent category. By characterizing CSO discharge volume and precipitation categorically, risk of AGI following the largest CSO/precipitation events are compared to risk of AGI following days without any events.

### Outcome Data

Statewide emergency department (ED) visit data for Massachusetts were obtained from the Center for Health Information and Analysis (CHIA), a state-level agency that collects and normalizes patient-level and financial data from all hospitals across Massachusetts. The Massachusetts Acute Hospital Case Mix Database contains data on ED visits as well as hospital inpatient discharges and outpatient observation stays for patients who originated in the ED.^37^ Data available for each visit include date, municipality of residence, patient age and sex, and primary and secondary diagnoses as defined by the International Classification of Diseases (ICD) 9^th^ or 10^th^ revision. Claims were de-identified and visits are assumed for analysis to represent separate individuals. Claims data were available from January 1, 2014 through September 30, 2019.

All individuals with AGI who entered a Massachusetts hospital through the ED during the study period, and whose permanent or temporary municipality of residence is in the study area are included. Cases of AGI were defined as any visit that included one or more of the following as a primary or secondary diagnosis: specified and unspecified infectious intestinal diseases (ICD-9: 001-009 or ICD-10: A00-A09), other and unspecified noninfectious gastroenteritis (ICD-9: 558.9 or ICD-10: K52.9), vomiting (ICD-9: 787.0, 787.01, 787.03 or ICD-10: R11.1, R11.10-R11.12, R11.2), and diarrhea (ICD-9: 787.91 or ICD-10: R19.7). Daily

AGI cases were aggregated to the municipal level based on residence for each study municipality over the study period. This study was designated as exempt from review by the Boston University Medical Campus Institutional Review Board (IRB number: H-42193).

### Statistical Analysis

A case time series design was used to assess associations between CSO/precipitation events and daily municipality-level cases of AGI. The case time series design is a self-matched method which allows for assessment of short-term risks of intermittent exposure events by comparing outcomes that occur at different times within individuals or small areas, such as municipalities.^38,39^ This approach allows for investigation of potentially lagged relationships and provides control for time-invariant characteristics by design, similar to other self-matched designs.^38,39^

The case time series model (Model 1) is specified as:

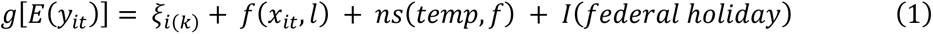

where *y*_*it*_ is the number of AGI cases in municipality *i* at time *t*. The baseline risks for different risk sets (e.g., temporal strata; *k=1,2,…ki*) are represented by *ξ*_*i*(*k*)_. The temporal strata used to define within-municipality comparison included year, month, and day of week. The function *f*(*x*_*it*_, *l*) specifies the association with the exposure in municipality *i* at time *t* along the lag dimension *l*. A uniform weighted lag structure was used that assumes that the probability of illness is constant across the lag period. The term *ns*(*temp*, *f*) describes a natural spline of average daily temperature with *f* degrees of freedom (three degrees of freedom was assumed). The temporal strata in the case time series model control for temporal trends across years, months, and days of the week, but *I*(*federal holiday*) is included as an indicator for public federal holidays because patterns of ED use may differ on holidays.^40,41^ Results generated by the case time series model include cumulative exposure-response relationships reported as cumulative risk ratio (CRR) as well as exposure-lag-response associations reported as daily risk ratio (DRR) over the lag period. CRR represents an adjusted estimate comparing the number of cases that occur over the lag period after an extreme event to the number of cases over the same lag period after days that are temporally matched by day of the week, month, and year when no event occurred.

Model 1 was used to estimate the association between extreme CSO events and AGI, and separately to estimate the association between extreme precipitation events and AGI (2 distinct models). Model 1 was also applied to the same two exposure metrics (CSO and precipitation events) stratified by municipality drinking water source: 1) exclusive use of the Merrimack River as a drinking water source, or 2) exclusive use of other drinking water sources (4 additional model iterations).

Precipitation is a driver of CSO events, and studies suggest that both precipitation and CSO events are independently associated with AGI.^32,42^ Model 2 includes extreme precipitation as a potential confounder in the association between CSO events and AGI:

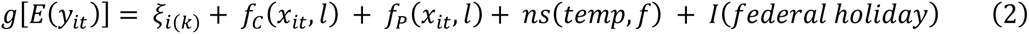

Model 2 components are the same as Model 1 except there are two functions—*f*_*C*_ (*x*_*it*_, *l*) and *f*_*P*_(*x*_*it*_, *l*)–that specify associations with extreme CSO discharge and precipitation events, respectively, in municipality *i* at time *t* along the lag dimension *l*. Model 2 was applied to a model for all municipalities in the study, as well as stratified groups by municipal drinking water source (3 models total). Analyses were conducted in R version 4.2.1 with version 2.4.7 of the *dlnm* package.

### Sensitivity Analysis

Models for all municipalities were tested with 4- and 7-day lag periods to consider different lagged relationships between exposure metrics and AGI. The definition of extreme CSO and precipitation events used in our categorical exposure variables was assessed by testing 90^th^, 95^th^, and 99^th^ percentile cutoffs. Three municipalities in the study area rely on the Merrimack River as a drinking water source intermittently, but data on how frequently or what proportion of drinking water needs are met by use of the Merrimack in these municipalities were not available, and the number of AGI cases from these municipalities was not sufficiently large for an independent analysis. The relationship between exposure events and AGI in municipalities that used any water from the Merrimack (exclusively and intermittently) was assessed as a sensitivity analysis. Finally, the association between individual municipality CSO discharge volumes and AGI in downstream municipalities was evaluated to account for bias that may be introduced by using a cumulative measure of upstream discharge.

## Results

### Descriptive Statistics

Over the course of the study period, CSS municipalities released a total of 3,012.6 million gallons (MG) of CSO discharge into the Merrimack River (Table S1). CSO events occurred in at least one CSS in the watershed on 447 days (21%) of the study period (January 1, 2014 – September 31, 2019). Both the frequency and size of CSO events varied among municipalities, with Manchester and Lowell contributing approximately 83% of total CSO discharge volume to the system. Cumulative daily upstream CSO discharge volume was highly variable during the study period, ranging from less than 0.1 MG to 189.0 MG with the 95^th^ percentile value at 27.3 MG and the 99^th^ percentile at 78.7 MG (Figure S2).

Precipitation events occurred in at least one of the municipalities in the study area on 972 days (46%) of the study period (Table S1). Precipitation events ranged from less than 0.1 inches of daily accumulation to 4.67 inches daily, with a 95^th^ percentile value of 1.24 inches and a 99^th^ percentile value of 1.97 inches. A positive correlation was observed between daily municipality-level precipitation and upstream cumulative CSO discharge on the day of the events (r=0.53) (Figure S3).

Massachusetts EDs documented 100,206 cases of AGI among residents of the study municipalities over the study period (Table 1). AGI patients were majority female (59.9%), and approximately 30% of patients were in age groups most susceptible to AGI (adults over 65 years old and children under 5 years old).^20^ The annual number of AGI ED visits increased by year over the study period, with 16,079 cases occurring in 2014 and 18,141 cases in 2018. Figure S4 shows the per capita rate of AGI among all study municipalities over the study period compared to the overall rate of AGI in the state of Massachusetts per the CHIA dataset.^37^

**Table 1:** Descriptive statistics for cases of acute gastrointestinal illness in 17 Massachusetts municipalities bordering the Merrimack River, January 1, 2014 – September 30, 2019.

| Characteristic | n (%) |
| --- | --- |
| Total number of AGI cases | 100,206 (100%) |
| Cases of AGI by year |  |
| 2014 | 16,079 (16.0%) |
| 2015 | 16,881 (16.8%) |
| 2016 | 16,903 (16.9%) |
| 2017 | 17,727 (17.7%) |
| 2018 | 18,141 (18.1%) |
| 2019 (though September 30) | 14,475 (14.4%) |
| Average number of cases of AGI per year | 17,427 |
| Cases of AGI by sex |  |
| Female | 59,989 (59.9%) |
| Cases of AGI by age group |  |
| < 5 years old | 14,845 (14.8%) |
| 5-19 years old | 15,094 (15.1%) |
| 20-64 years old | 53,999 (53.9%) |
| 65+ years old | 16,268 (16.2%) |
| Cases of AGI by municipal drinking water source |  |
| Merrimack River exclusively (4 municipalities) | 64,278 (64.2%) |
| Merrimack River intermittently (3 municipalities) | 7,649 (7.6%) |
| Other sources (10 municipalities) | 28,279 (28.2%) |

### Model Results

#### All municipalities

In models adjusted for temporal trends and temperature but not adjusted for precipitation, the cumulative risk ratio (CRR) of AGI was 22% higher (CRR: 1.22 [95% CI: 1.05, 1.42]) in the four days following 95^th^ and 63% higher (1.63 [1.07, 2.47]) in the four days following 99^th^ percentile CSO events compared to days with no CSO events when considering all municipalities (Figure 2a). A marginal association that was not statistically significant was observed between 90^th^ percentile CSO events and AGI (1.07 [0.96, 1.19]). No increase in risk was observed following CSO events below the 90^th^ percentile. These associations remain largely unchanged among all municipalities after adjusting for precipitation, with a 17% increase in CRR of AGI in the 4 days after 95^th^ percentile CSO events (1.17 [0.98, 1.39]) and a 62% increase after 99^th^ percentile CSO events (1.62 [1.04, 2.51]) (Figure 2b). Extreme precipitation is also associated with a 13% increase in CRR of AGI in the 4 days following 95^th^ percentile precipitation events (1.13 [1.00, 1.27]), and a marginal increase following 99^th^ percentile precipitation events (1.07 [0.82, 1.41]) (Figure 2c). Results of all models, including those that assessed the association between extreme events and AGI among municipalities that use any water from the Merrimack and those that used a seven-day lag period, are included in Tables S2 and S3.

**Figure 2:**
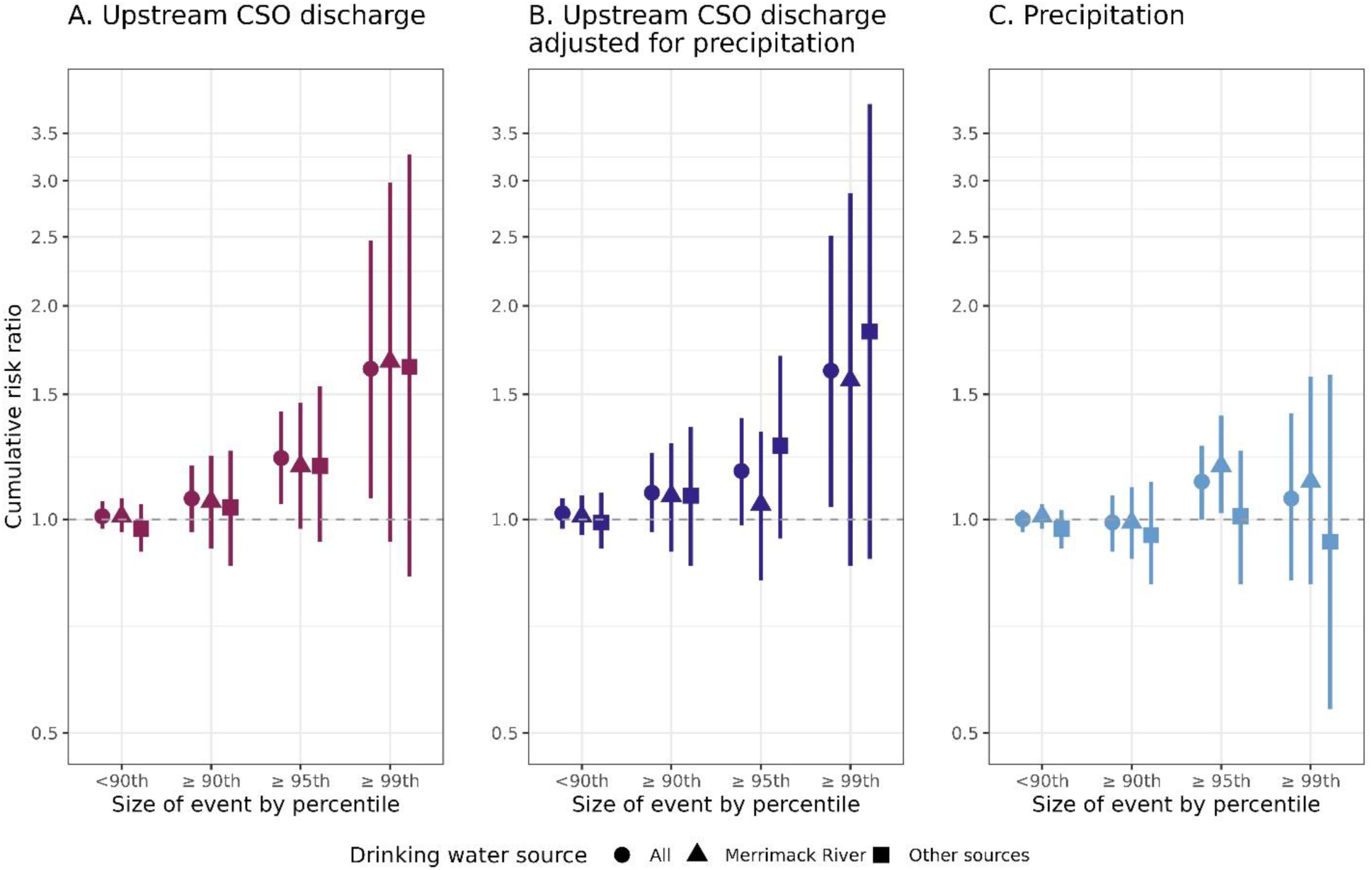
Cumulative risk ratio (CRR) of acute gastrointestinal illness (AGI) over the four days following A) combined sewer overflow (CSO) events, B) CSO events adjusting for precipitation, and C) precipitation events. Model results are shown for all municipalities and by drinking water source (Merrimack River or other sources), as well as events of different sizes: <90^th^ percentile, ≥90^th^ percentile, ≥95^th^ percentile, and ≥99^th^ percentile events. Municipalities that intermittently use the Merrimack River as a drinking water source were included in the analysis for all municipalities but not in the drinking water stratification analysis.

#### Stratification by drinking water source

We did not observe a difference in the unadjusted CRR of AGI in municipalities with different drinking water sources following 95^th^ percentile CSO events after stratification by drinking water source (Merrimack River: 1.19 [0.97, 1.46]; Other sources: 1.19 [0.93, 1.54]). After adjusting for precipitation, CRR of AGI was only marginally elevated following 95^th^ percentile CSO events among municipalities that exclusively get their drinking water from the river (1.05 [0.82, 1.33]), while the CRR of AGI among municipalities that do not use the Merrimack River as a drinking water source was slightly higher (1.27 [0.94, 1.70]) (Figure 2b). CRR of AGI was elevated regardless of drinking water source after 99^th^ percentile CSO events compared to days with no CSO events (Merrimack River: 1.57 [0.86, 2.88]; Other sources: 1.84 [0.88, 3.85]) (Figure 2b).

#### Daily relative risk

Comparison of daily risk ratios (DRR) of AGI in municipalities with different drinking water sources over the four-day lag period shows that the magnitude of the strongest observed increase in DRR was approximately the same between drinking water groups (Merrimack River: 1.36 [1.05, 1.76]; Other sources: 1.45 [1.07, 1.96]), but occurred on different days within that period (Figure 3). The strongest association between extreme CSO events and AGI was observed two days after an event among municipalities that use the Merrimack as their exclusive drinking water source, and one day after an event among municipalities with other drinking water sources. A similar temporal pattern was observed in the four days following 95^th^ percentile CSO events, though the magnitude of the associations were attenuated (Figure S5).

**Figure 3:**
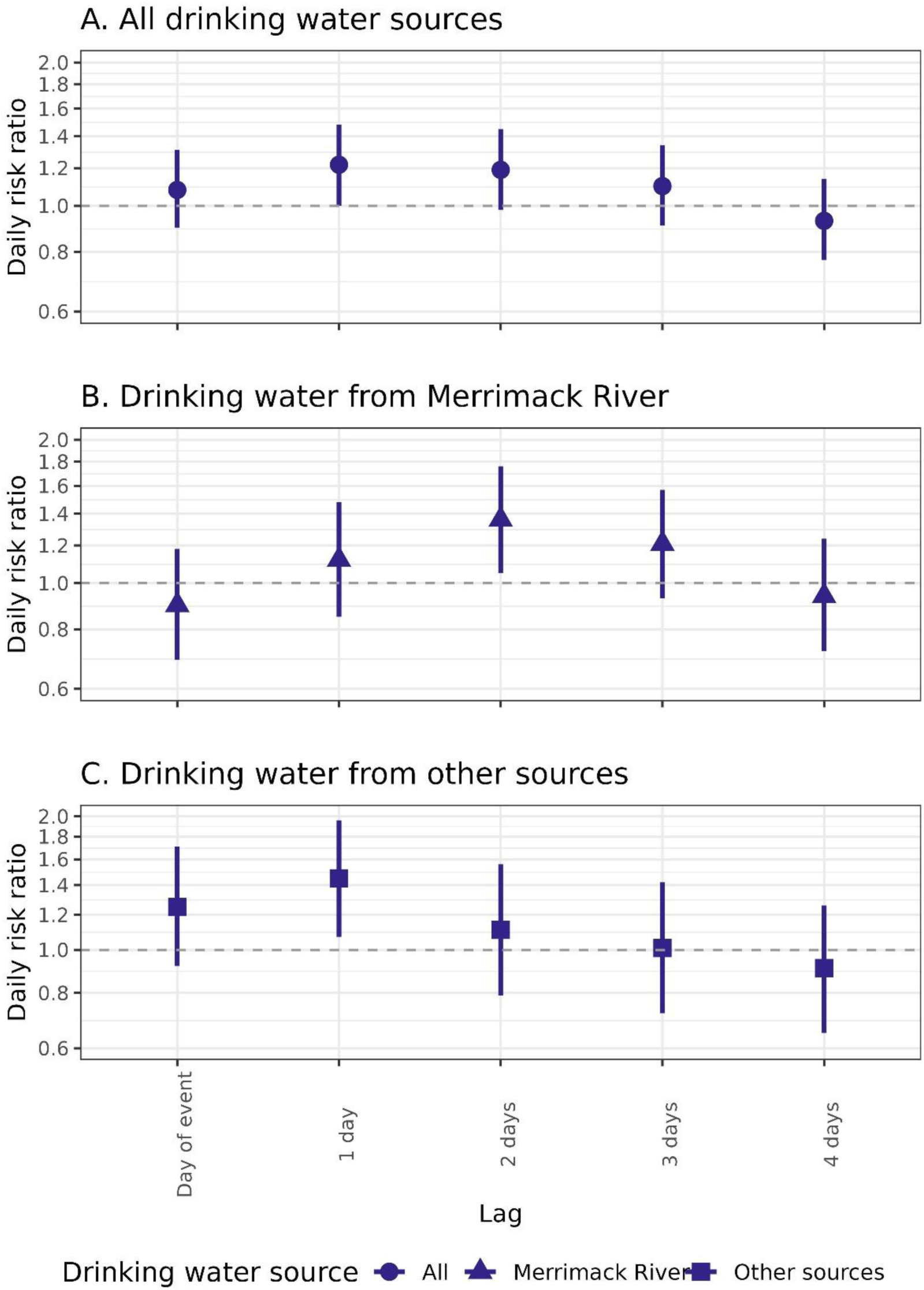
Daily risk ratio (DRR) of acute gastrointestinal illness (AGI) over a 4-day lag period following of 99^th^ percentile combined sewer overflow (CSO) events and adjusted for precipitation in A) municipalities with all drinking water sources, B) municipalities that exclusively get their drinking water from the Merrimack River, and C) municipalities that exclusively get their drinking water from sources other than the Merrimack. Municipalities that intermittently use the Merrimack River as a drinking water source were included in the analysis for all municipalities but not in the drinking water stratification analysis.

#### Sensitivity analyses

Results from the sensitivity analysis assessing the relationship between CSO events from individual CSS municipalities and cases of AGI in downstream municipalities are shown in Table S4. Elevated CRR of AGI is observed among downstream municipalities 4-days after CSO events in Manchester (1.09 [0.93, 1.29]) and Lowell (1.16 [0.95, 1.42]), though neither of these associations are statistically significant.

## Discussion

In this analysis of the association between CSO events and ED visits for AGI, we observed higher risk of AGI in the 4 days following extreme CSO events compared to days with no CSO events. Results were robust to adjustment for potential confounding by precipitation. The magnitude of CSO discharge volume was an important factor in the association between CSO events and AGI, with the largest associations observed following ≥95^th^ and ≥99^th^ percentile CSO events. Stratification by drinking water source (Merrimack River or other source) suggests that the association between ≥95^th^ percentile CSO events and AGI after adjusting for extreme precipitation events was slightly more pronounced in municipalities that do not rely on the Merrimack River as a drinking water source, though CRR of AGI was elevated after the largest CSO events (>99^th^ percentile) regardless of drinking water source. The temporal pattern of daily risk in the 4 days following 99^th^ percentile CSO events differed between municipalities that do and do not use the Merrimack River as a drinking water source, which may suggest different exposure pathways.

The magnitude of the associations observed in this analysis are similar to or larger than those found in previous studies, though methodological differences and study populations make direct comparisons difficult. Using a similar self-matched study design, Brokamp et al. found a 16% higher odds of AGI after any CSO event among children living within 500m of a CSO outfall (OR: 1.16 [1.04, 1.30]).^33^ This study differed in notable ways from our study, including in the approach to exposure classification, study population, and the inclusion of CSO volume and precipitation, but the magnitude of the association was similar to the CRR we observed following 95^th^ percentile CSO events. On the other hand, Miller et al. did not use a self-matched study design and found a smaller but more precise association between large CSO events and AGI after adjusting for precipitation among Atlanta residents of all ages (rate ratio: 1.09 [1.03, 1.14]).^32^ The definition of a large CSO event in Miller et al. was ≥56.6 MG of discharge, which falls in the range between 95^th^ (27.3 MG) and 99^th^ percentile (78.7 MG) CSO discharge events in our study. In an analysis in the Merrimack Valley region of Massachusetts that used extreme precipitation as a proxy for CSO events, Jagai et al. found a 13% increase in cumulative risk of AGI following extreme precipitation events (equivalent to 1.97 in., CRR: 1.13 [1.00, 1.28]).^31^ This result is remarkably similar to the association observed in this analysis between 95^th^ percentile precipitation events (equivalent to 1.24 in.) and cumulative risk of AGI over four days observed in our study. Overall, we find that the strength of the associations observed in this study between 95^th^ percentile CSO or precipitation events and AGI are fairly consistent with previous findings, but that the CRR of AGI following 99^th^ percentile CSO events is notably higher than what has been reported in similar studies.

Our findings suggest that a nonlinear dose-response relationship exists between upstream CSO discharge volume and AGI, a finding consistent with previous studies.^31,32^ Miller et al. similarly found that only large CSO events (≥75^th^ percentile CSO events by volume, equivalent to 56.5 MG) were significantly associated with AGI in the week following an event, but models estimating the association between any CSO event and AGI resulted in no association.^32^ Jagai et al. found that only the most extreme precipitation events (99^th^ percentile, equivalent to 1.97 in) were significantly associated with increased cumulative risk of AGI; lower magnitude, not statistically significant associations were found for lower percentile precipitation events.^31^ These results are particularly relevant as the frequency and intensity of extreme precipitation events (and extreme CSO events as a result) are expected to increase due to climate change in the Northeast and Midwest where most CSS in the US are located.^3^

The results of this analysis suggest that there is an increase in risk of AGI following 99^th^ percentile CSO events in municipalities that source their drinking water from the Merrimack River that could be related to contaminated drinking water, though these results should be interpreted with caution. The later peak in DRR of AGI following extreme CSO events in municipalities that get their drinking water from the Merrimack compared to those that do not could also suggest a longer lag due to additional time for source water intake, treatment, and distribution. However, extreme precipitation events were associated with AGI most prominently in municipalities that use the Merrimack River as their exclusive or intermittent drinking water source (Table S2C), which could instead suggest contamination of drinking water due to pathogen infiltration or cross contamination of leaky sewage and water distribution pipes.^43,44^ The large confidence intervals and lack of statistical significance among any of the stratified drinking water groups in this study, possibly due to smaller case counts, further limits the conclusions that can be made about the potential for drinking water contamination.

The evidence in the literature is inconclusive on the potential for drinking water contamination after CSO events. While it is conceptually possible for treated drinking water to be contaminated by CSO discharge,^45,46^ studies that incorporated drinking water source contamination following discharge events either did not include CSO event data in the exposure^30,31^ or found that controlling for CSO events as a binary covariate did not alter the association between precipitation and AGI.^42^ We are also unaware of an accounting of how many municipalities have a drinking water source that is impacted by CSO discharge, so it is unclear how widespread this issue might be, though the results of this analysis suggest that drinking water contamination may not be the only important exposure pathway in a CSO- impaired river system. Understanding the critical exposure pathway(s) following discharge events is an important direction for future work that has direct implications for infrastructure investment decisions and policies designed to protect public health.

The findings of this study suggest that CSO events and AGI were more strongly associated across the four days following an event compared to a longer lag period of seven days. These findings are consistent with previous studies that found significant associations between sewage discharge events and AGI over lag periods from 2-8 days.^30–33^ A four-day lag period is also consistent with the incubation times of some of the pathogens found in waterways contaminated with sewage.^1,9,47^ Some AGI-causing viral and bacterial pathogens have incubation periods that are consistent with the short lag identified in this study including noroviruses (12-48 hours), rotavirus (1-3 days), *Shigella* (12 hours-7 days), and enterotoxigenic *E. coli* (12-72 hours).^47^ Viral waterborne pathogens can be infectious at low doses, persistent in environmental media, and less effectively removed by conventional drinking water treatment compared to bacteria.^48,49^ Another consideration with a short lag time is time between CSO event and exposure, which could differ based on exposure pathway. It is possible that people exposed primarily through recreational activity or aerosolized pathogens could ingest contaminated water within hours after a CSO event, though our analysis was not spatially refined enough to assess the possibility of exposure through the latter pathway.

This study has a number of potential limitations. First, the analysis is limited by a lack of individual-level information on exposure, including rates of consumption or quality of drinking water, amount or timing of contact with recreational waters, and/or recreational surface water quality experienced by individuals within the study population. The lack of individual exposure data limits our ability to gain insights about specific exposure pathways. Moreover, lack of information about local water contamination and potential local responses to CSO events (e.g., adjustments to drinking water treatment processes), may have led to substantial exposure misclassification and likely biased results towards the null hypothesis of no association. Second, the AGI case data were deidentified, precluding identification of patients that may have been treated for AGI more than once. Our inability to account for the correlation among repeated episodes within an individual may have resulted in confidence intervals that are perhaps too narrow. Third, people seeking care for AGI in the ED likely represent the most severe cases of gastrointestinal illness, given that 10% or fewer of all cases of AGI come to medical attention in acute care settings.^50^ Thus, it is unclear whether these results are generalizable to milder episodes of AGI or to other communities across the country.

On the other hand, this study has some notable strengths. First, the case time series design minimizes potential confounding by differences between municipalities and effectively controls for temporal trends. Second, by using daily measures of CSO discharge volume instead of approaches taken in other studies to measure CSO exposure, such as precipitation as a proxy for CSO events,^31^ modeling CSO events as binary exposures,^33^ or aggregating to a weekly time step,^32^ this study more directly evaluates the relationship between CSO discharge and health. Our use of daily CSO and precipitation data allowed for a refined temporal analysis in the days following an exposure event, and CSO volume data provided the opportunity to assess potential dose-response relationships between CSO discharge and AGI. We also considered the spatial relationship between CSS municipalities and downstream municipalities in our approach to assigning exposure. While there are limitations to characterization of exposure in this study by considering upstream discharge, this approach is representative of how CSO discharge moves through the area and accumulates in downstream stretches of the river. The PRISM dataset provided spatial resolution of precipitation estimates that may account for heterogeneity in rainfall that is not captured by a regional weather station, the latter of which has been the source for precipitation data in other studies.^31,32^ Finally, reliance on the Merrimack River as a drinking water source downstream of CSO outfalls provided an opportunity to compare associations between municipalities that do and do not get their drinking water from this CSO-impacted source. To our knowledge, only three studies report the association between sewage-related discharge events and health outcomes in regions with modern wastewater infrastructure where drinking water intakes exist downstream of outfalls.^30,31,42^ In those studies, the exposure was either extreme rainfall as a proxy for CSO events,^31^ undertreated sewage discharge events,^30^ or precipitation with CSO discharge included as a covariate.^42^

Our findings suggest that extreme CSO events increase the risk of ED visits for AGI in downstream municipalities in the four days following an event, and that a non-linear dose-response relationship exists between CSO discharge and ED visits for AGI. The largest CSO events are associated with AGI across drinking water sources and concurrent precipitation levels, although we observed differences in the timing and magnitude of associations based on drinking water source. Although our study is ecological in design, our findings indicate that CSS may negatively impact public health. As precipitation events intensify due to climate change, large-volume CSO events may become more common and the risk of AGI in downstream municipalities may increase.

## Data Availability

Health outcome data were made available to the authors by the Center for Health Information and Analysis under a data use agreement and cannot be shared by the authors. Exposure data that originated from public sources are available from the corresponding author upon reasonable request.

## Acknowledgements

The authors acknowledge staff at US EPA Region 1 and the City of Manchester for their help locating CSO discharge data. We are also grateful to Kevin Brander of the Massachusetts Department of Environmental Protection for sharing his expertise and knowledge of the study system and to Aine Studdert-Kennedy for her assistance with data entry.

