## Supplemental materials for "Association between combined sewer overflow events and gastrointestinal illness in Massachusetts municipalities with and without river-sourced drinking water, 2014-2019"

### Supplemental Material

Beth M. Haley, Yuantong Sun, Jyotsna S. Jagai, Jessica H. Leibler, Robinson Fulweiler, Jacqueline Ashmore, Gregory A. Wellenius, Wendy Heiger-Bernays

Tables

**Table S1: Descriptive statistics for exposure metrics (January 1, 2014 – September 30, 2019)**

| | CSO Events | Precipitation Events[a] |
|---|---|---|
| Total days with event in watershed (% days) | 447 (21%) | 972 (46%) |
| Exposure distribution and percentile cutoffs[b] | | |
| Range | <0.1 – 189.0 MG | <0.1 - 4.67 in |
| 90th percentile | 15.4 MG | 0.88 in |
| 95th percentile | 27.3 MG | 1.24 in |
| 99th percentile | 78.7 MG | 1.97 in |
| Total days with discharge events by municipality (% days) | | |
| Manchester | 332 (16%) | - |
| Nashua | 121 (6%) | - |
| Lowell | 161 (8%) | - |
| Lawrence | 53 (3%) | - |
| Haverhill | 222 (11%) | - |
| Total discharge (MG) in watershed (annual average) | 3,012.6 (523.9) | - |
| Manchester | 1,379.0 (239.8) | - |
| Nashua | 156.0 (27.1) | - |
| Lowell | 1,113.6 (193.7) | - |
| Lawrence | 194.0 (33.7) | - |
| Haverhill | 170.0 (29.6) | - |

[a]Precipitation exposure assigned by municipality (n=17)
[b]CSO event percentiles represent percentile cutoffs for upstream cumulative CSO discharge (MG)

**Table S2: Cumulative risk ratio (CRR) of AGI over a 4-day lag period following A) CSO events, B) CSO events adjusted for precipitation, and C) precipitation events.**

| **A) CSO events (unadjusted)** | | | | |
|---|---|---|---|---|
| Lag period | 4-day lag | | | |
| Size of event by percentile | All municipalities | Merrimack River source exclusively | Merrimack River source ever | Other drinking water sources exclusively |
| <90th | 1.01 (0.97, 1.06) | 1.01 (0.96, 1.07) | 1.03 (0.98, 1.08) | 0.97 (0.90, 1.05) |
| ≥90th | 1.07 (0.96, 1.19) | 1.06 (0.91, 1.23) | 1.08 (0.94, 1.24) | 1.04 (0.86, 1.25) |
| ≥95th | 1.22 (1.05, 1.42)* | 1.19 (0.97, 1.46) | 1.23 (1.02, 1.49)* | 1.19 (0.93, 1.54) |
| ≥99th | 1.63 (1.07, 2.47)* | 1.67 (0.93, 2.98) | 1.58 (0.93, 2.69) | 1.64 (0.83, 3.27) |
| **B) CSO events (adjusted for precipitation)** | | | | |
| Lag period | 4-day lag | | | |
| Size of event by percentile | All municipalities | Merrimack River source exclusively | Merrimack River source ever | Other drinking water sources exclusively |
| <90th | 1.02 (0.97, 1.07) | 1.01 (0.95, 1.08) | 1.03 (0.97, 1.10) | 0.99 (0.91, 1.09) |
| ≥90th | 1.09 (0.96, 1.24) | 1.08 (0.90, 1.28) | 1.10 (0.93, 1.29) | 1.08 (0.86, 1.35) |
| ≥95th | 1.17 (0.98, 1.39) | 1.05 (0.82, 1.33) | 1.12 (0.90, 1.40) | 1.27 (0.94, 1.70) |
| ≥99th | 1.62 (1.04, 2.51)* | 1.57 (0.86, 2.88) | 1.52 (0.87, 2.66) | 1.84 (0.88, 3.85) |
| **C) Precipitation events** | | | | |
| Lag period | 4-day lag | | | |
| Size of event by percentile | All municipalities | Merrimack River source exclusively | Merrimack River source ever | Other drinking water sources exclusively |
| <90th | 1.00 (0.96, 1.03) | 1.01 (0.97, 1.05) | 1.01 (0.97, 1.05) | 0.97 (0.91, 1.03) |
| ≥90th | 0.99 (0.90, 1.08) | 0.99 (0.88, 1.11) | 1.00 (0.90, 1.12) | 0.95 (0.81, 1.13) |
| ≥95th | 1.13 (1.00, 1.27)* | 1.19 (1.02, 1.40)* | 1.18 (1.02, 1.37)* | 1.01 (0.81, 1.25) |
| ≥99th | 1.07 (0.82, 1.41) | 1.13 (0.81, 1.59) | 1.10 (0.80, 1.51) | 0.93 (0.54, 1.60) |

In all cases, the reference category is days with no CSO/precipitation events. Results are shown for all municipalities and by drinking water source (Merrimack River exclusively, any use of the Merrimack, or exclusively other sources), as well as events of different sizes: <90th percentile, ≥90th percentile, ≥95th percentile, and ≥99th percentile events. Statistical significance is indicated by an asterisk (*).

**Table S3: Cumulative risk ratio (CRR) of AGI over a 7-day lag period following A) CSO events, B) CSO events adjusted for precipitation, and C) precipitation events.**

| **A) CSO events (unadjusted)** | | | | |
|---|---|---|---|---|
| Lag period | 7-day lag | | | |
| Size of event by percentile | All municipalities | Merrimack River source exclusively | Merrimack River source ever | Other drinking water sources exclusively |
| $<90^{th}$ | 1.03 (0.97, 1.09) | 1.03 (0.96, 1.11) | 1.04 (0.98, 1.12) | 0.99 (0.89, 1.10) |
| $\geq 90^{th}$ | 0.94 (0.82, 1.09) | 0.92 (0.76, 1.11) | 0.96 (0.81, 1.15) | 0.91 (0.71, 1.15) |
| $\geq 95^{th}$ | 1.10 (0.91, 1.34) | 1.01 (0.77, 1.31) | 1.09 (0.85, 1.39) | 1.12 (0.81, 1.55) |
| $\geq 99^{th}$ | 1.46 (0.84, 2.55) | 1.49 (0.69, 3.22) | 1.49 (0.73, 3.01) | 1.32 (0.52, 3.36) |
| **B) CSO events (adjusted for precipitation)** | | | | |
| Lag period | 7-day lag | | | |
| Size of event by percentile | All municipalities | Merrimack River source exclusively | Merrimack River source ever | Other drinking water sources exclusively |
| $<90^{th}$ | 1.06 (0.99, 1.13) | 1.06 (0.98, 1.16) | 1.08 (1.00, 1.17)* | 1.00 (0.89, 1.13) |
| $\geq 90^{th}$ | 1.04 (0.88, 1.23) | 1.03 (0.82, 1.29) | 1.08 (0.88, 1.33) | 0.96 (0.72, 1.29) |
| $\geq 95^{th}$ | 1.12 (0.89, 1.41) | 0.96 (0.71, 1.32) | 1.05 (0.79, 1.40) | 1.27 (0.86, 1.86) |
| $\geq 99^{th}$ | 1.70 (0.95, 3.06) | 1.75 (0.79, 3.88) | 1.70 (0.81, 3.55) | 1.66 (0.61, 4.52) |
| **C) Precipitation events** | | | | |
| Lag period | 7-day lag | | | |
| Size of event by percentile | All municipalities | Merrimack River source exclusively | Merrimack River source ever | Other drinking water sources exclusively |
| $<90^{th}$ | 1.01 (0.97, 1.06) | 1.02 (0.96, 1.08) | 1.02 (0.96, 1.07) | 1.01 (0.93, 1.09) |
| $\geq 90^{th}$ | 0.89 (0.79, 1.00) | 0.87 (0.75, 1.02) | 0.89 (0.77, 1.03) | 0.88 (0.70, 1.10) |
| $\geq 95^{th}$ | 1.03 (0.88, 1.20) | 1.06 (0.86, 1.30) | 1.09 (0.90, 1.32) | 0.89 (0.66, 1.18) |
| $\geq 99^{th}$ | 0.77 (0.54, 1.10) | 0.72 (0.46, 1.12) | 0.78 (0.51, 1.19) | 0.69 (0.34, 1.38) |

In all cases, the reference category is days with no CSO/precipitation events. Results are shown for all municipalities and by drinking water source (Merrimack River exclusively, any use of the Merrimack, or exclusively other sources), as well as events of different sizes: $<90^{th}$ percentile, $\geq 90^{th}$ percentile, $\geq 95^{th}$ percentile, and $\geq 99^{th}$ percentile events. Statistical significance is indicated by an asterisk (*).

**Table S4: Cumulative risk ratio (CRR) of AGI in downstream municipalities over a 4-day lag period following 95th percentile CSO events occurring in specific CSS municipalities**.

| | Number of ≥95th percentile CSO events | All municipalities | Merrimack River source exclusively | Merrimack River source ever | Other drinking water sources exclusively |
|---|---|---|---|---|---|
| Manchester | 19 | 1.09 (0.93, 1.29) | 1.05 (0.85, 1.30) | 1.06 (0.87, 1.29) | 1.19 (0.88, 1.60) |
| Nashua | 3 | 0.80 (0.52, 1.23) | 0.79 (0.46, 1.38) | 0.80 (0.48, 1.35) | 0.77 (0.34, 1.74) |
| Lowell | 18 | 1.16 (0.95, 1.42) | 1.18 (0.89, 1.55) | 1.19 (0.92, 1.53) | 1.11 (0.80, 1.55) |
| Lawrence | 4 | 0.92 (0.42, 2.00) | - | - | - |
| Haverhill | 1 | 0.13 (0.01, 2.58) | - | - | - |

When considered as individual events, a 95th percentile CSO event is equivalent to discharge of 14.53 MG or larger from one CSS municipality in one day. For those CSS municipalities located upstream of drinking water intakes, the results of a stratified analysis by drinking water source are also shown.

Figures

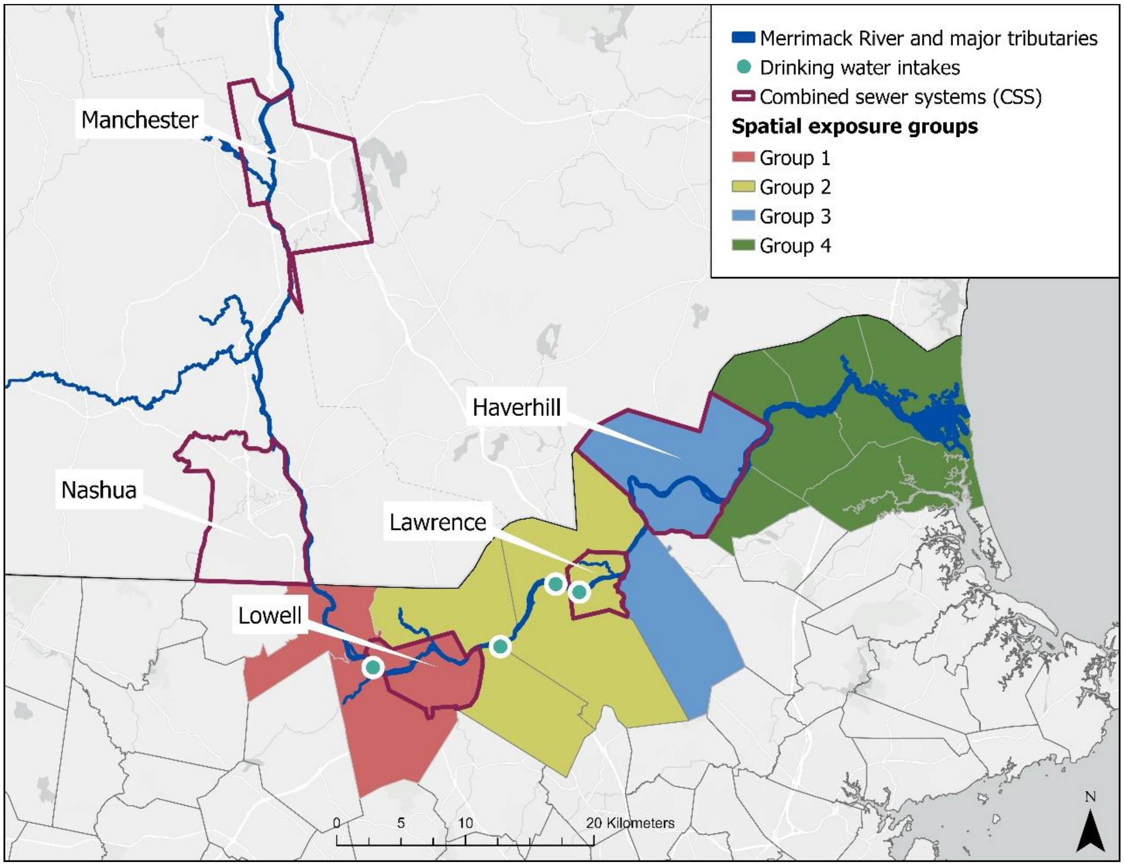


**Figure S1: Map of spatial groups used to characterize CSO exposures.**

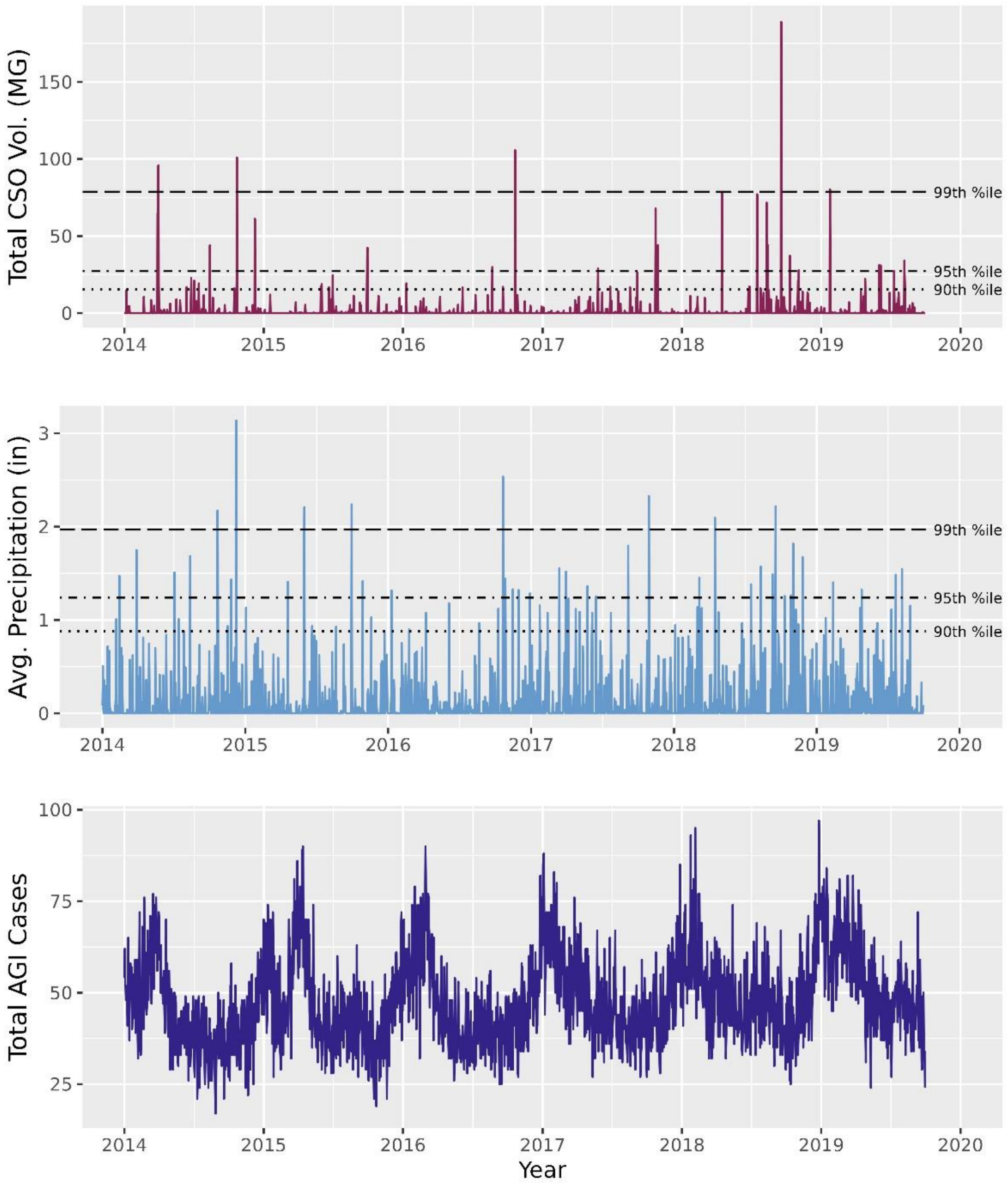


**Figure S2: Time series of daily cumulative CSO discharge, average precipitation, and total AGI cases in study area (January 2014 – September 2019).** The time series for CSO discharge shows total daily discharge in the watershed; precipitation time series is a daily average of the total precipitation in all study municipalities. The 90$^{th}$, 95$^{th}$, and 99$^{th}$ percentile designations are indicated for cumulative upstream CSO discharge volume and total daily precipitation.

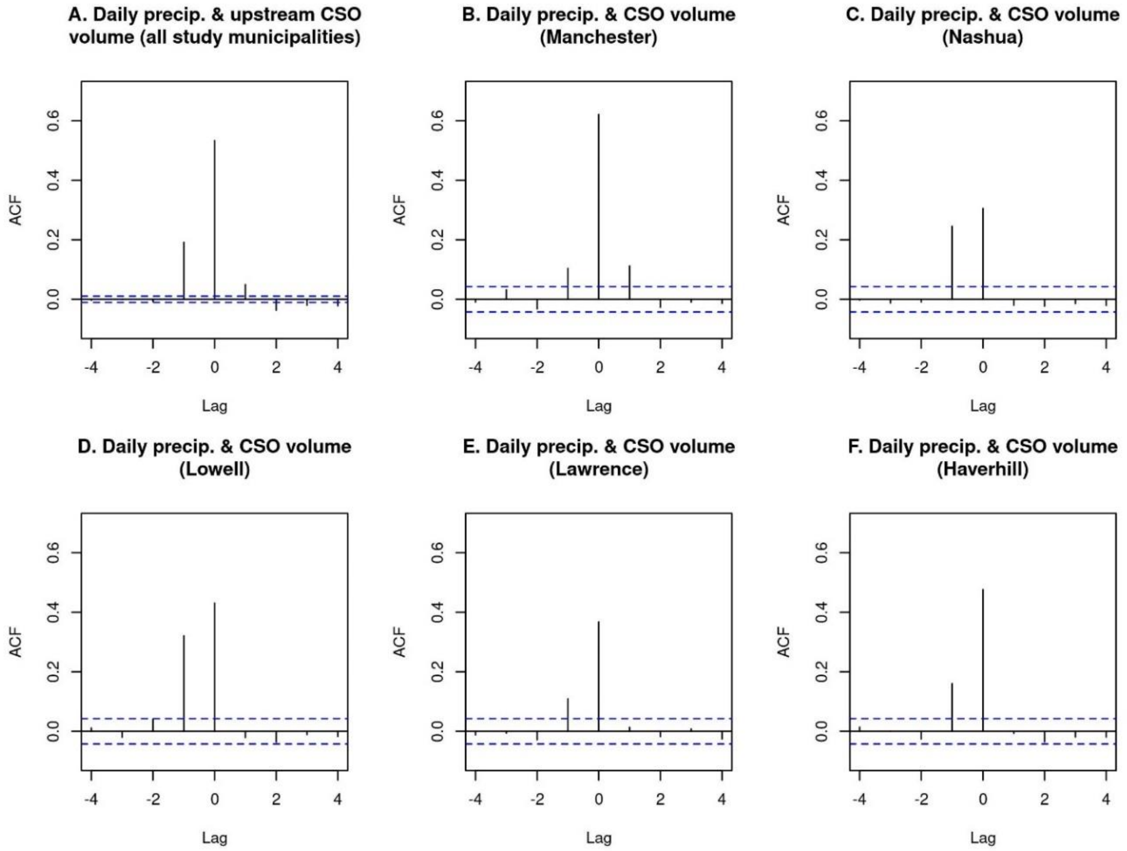


**Figure S3: Cross-correlations between daily precipitation and CSO discharge volume.** Figure S3a shows the cross-correlation between daily municipality-level precipitation and cumulative upstream CSO discharge volume in all study municipalities, whereas Figures S3b-f show the cross-correlations between daily precipitation and CSO discharge volume in the 5 CSS municipalities included in the study. If a significant correlation (indicated by a vertical line extending beyond the dashed horizontal confidence interval lines) occurs on lag day 0 (h=0), the events are correlated on the same day that they occur. If there is significant correlation on a negative lag day (meaning to the left of zero, or -h), precipitation leads CSO discharge, meaning that precipitation is predictive of CSO discharge occuring h days later. If there is a significant correlation on a positive lag day (meaning to the right of zero, or h), precipitation lags behind CSO discharge, meaning that above average CSO discharge is likely to lead to above average precipitation the next day.

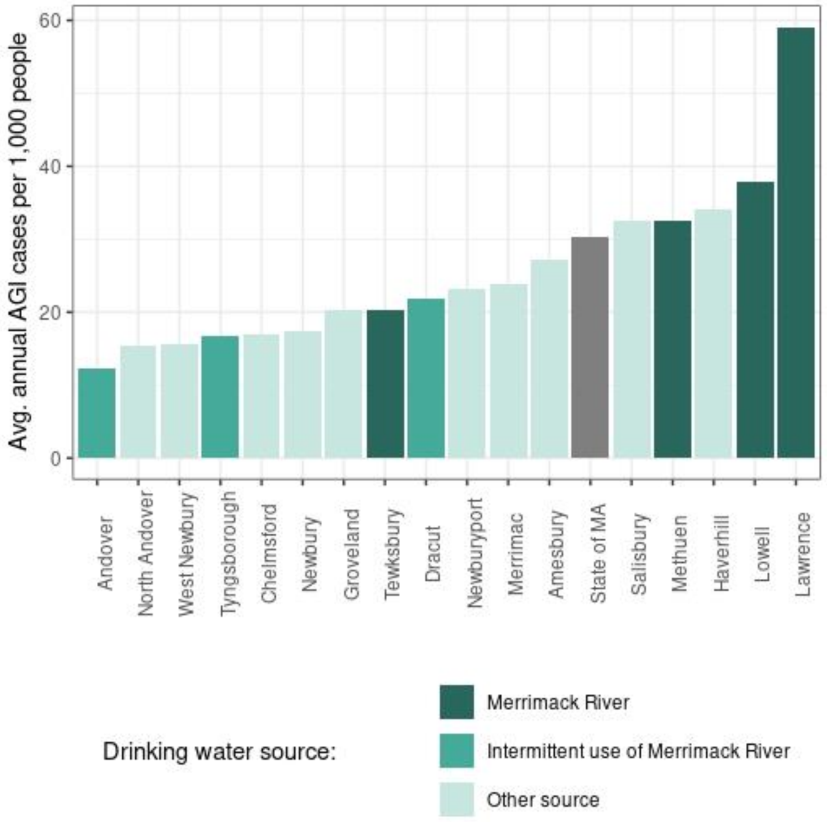


**Figure S4: Average annual cases of AGI per thousand people among study municipalities compared to background levels of AGI in the state of Massachusetts**. Annual population estimates for the state of MA and study municipalities were accessed through the American Community Survey 5-year estimates[1] and City and Town Population Totals[2] from the US Census.

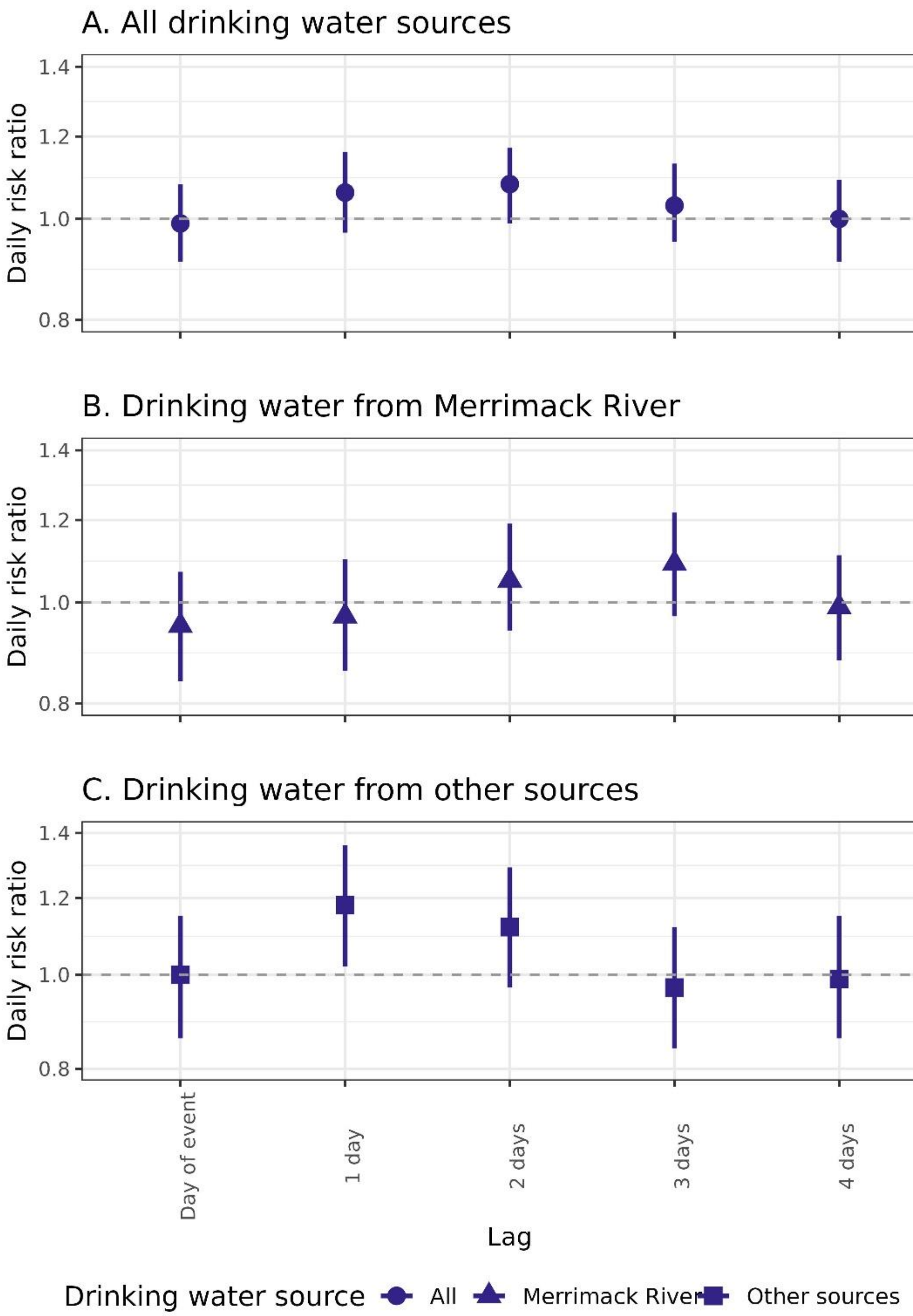


**Figure S5: Daily risk ratio (DRR) of AGI over a 4-day lag period after 95$^{th}$ percentile CSO events adjusted for 95$^{th}$ percentile precipitation events** among A) all study municipalities, B) municipalities that exclusively source their drinking water from the Merrimack River, and C) municipalities that exclusively get their drinking water from other sources.
